# Telecommuting frequency and preference among Japanese workers according to regional cumulative COVID-19 incidence: a cross-sectional study

**DOI:** 10.1101/2021.05.18.21257423

**Authors:** Hiroka Baba, Kazunori Ikegami, Hajime Ando, Hisashi Eguchi, Mayumi Tsuji, Seiichiro Tateishi, Masako Nagata, Shinya Matsuda, Yoshihisa Fujino, for the CORoNaWork Project

## Abstract

This study aimed to examine the relationship between telecommuting and the regional cumulative COVID-19 incidence. This was a cross-sectional study analyzing 13,468 office workers. The participant groups, according to the level of cumulative COVID-19 incidence by prefecture, were used as the predictor variable, and telecommuting frequency and preference were used as outcomes. We employed an ordinal logistic regression analysis. In regions with a high cumulative COVID-19 incidence, the proportion of participants who telecommuted more than two days per week was 34.7%, which was approximately 20% higher than in other regions. Telecommuting preference was stronger in areas with higher COVID-19 influence. However, in other regions, the proportion of participants who did not want to telecommute was higher than that of those who wanted to telecommute. We found that telecommuting frequency and preference were higher in regions with high cumulative COVID-19 incidence.

## Introduction

At the end of 2019, an outbreak of pneumonia of unknown cause was confirmed in Wuhan, People’s Republic of China, and a novel coronavirus (SARS-CoV-2) was discovered as the etiologic agent (Lai et al., 2020). The communicable disease caused by SARS-CoV-2 has been designated as Coronavirus disease 2019 (COVID-19), and this pandemic continued in 2021. In Japan, the first, second, and third waves of the COVID-19 pandemic occurred in April 2020, July to August 2020, and December 2020 to February 2021, respectively.

The COVID-19 pandemic greatly impacted the work environment and practices, resulting in changes in work systems and management (Alon et al., 2020). Globally, the expansion of telecommuting in the wake of the COVID-19 pandemic has been one of the most drastic changes in the way people work in recent decades (Belzunegui-Eraso & Erro-Garcés, 2020; Khan & Javed Hasan, 2020). In February 2020, the Japanese government released a basic policy describing preventive measures for the COVID-19 epidemic; this included a recommendation for companies to implement telecommuting to prevent the spread of COVID-19 (Ministry of Health, Labor and Welfare, 2020). As the COVID-19 incidence continued to increase throughout Japan, more companies introduced telecommuting, and the proportion of telecommuters increased. A survey conducted by the Tokyo Metropolitan Government at the end of February 2021, among companies with 30 or more employees, reported that more than 50% of companies have introduced telecommuting systems (Tokyo Metropolitan Government, 2021). In a survey conducted in April, May, and November 2020 among approximately 20,000 workers in the Japanese private sector, the implementation rate of telecommuting was approximately 25% (PERSOL RESEARCH AND CONSULTING Co., 2020a, 2020b, 2020c). However, the survey also reported regional differences in the status of telecommuting implementation, with the highest and lowest proportions of telecommuters being 45.8% and 3.5%, respectively, in November 2020 (PERSOL RESEARCH AND CONSULTING Co., 2020b).

Many workers could have recognized that workplace and commuting have high risk for COVID-19 infection and that telecommuting is an important measure to prevent COVID-19. The regional state of COVID-19 epidemic may reflect discrepancies in the spread of telecommuting. Its introduction is thought to be mainly for preventing increased COVID-19 incidence, and the pandemic may be suppressed in regions with a high proportion of telecommuters. There could also be regional discrepancies in the telecommuting preference. Workers who live in a region with high COVID-19 incidence may have higher telecommuting preference, or there may be a contradiction that workers in the areas where telecommuting is not spreading have a higher telecommuting preference. We consider it necessary to evaluate the relationship between the degree of cumulative COVID-19 incidence in the region and the telecommuting frequency, or telecommuting preference. This study clarified the relationship between telecommuting and the state of the COVID-19 pandemic in these regions.

## Methods

### Study Design and Setting

We conducted a prospective cohort study of the novel coronavirus and work study (CORoNaWork study) through a research group consisting of the University of Occupational and Environmental Health, using collaborative online research. Data were obtained through a self-administered questionnaire survey conducted by a Japanese Internet survey company (Cross Marketing Inc. Tokyo); a baseline survey was conducted from December 22 to 25, 2020. Incidentally, during the baseline survey, the number of COVID-19 infections and deaths were overwhelmingly higher than in the first and second waves; therefore, Japan was on maximum alert during this third wave. This study implemented a cross-sectional design using part of a baseline survey of the CORoNaWork study. Fujino et al. introduced the details of this study protocol (Fujino et al., 2021).

### Participants

Participants in this survey were aged between 20 and 65 years and working at the time of the baseline survey. A total of 33,087 participants, who were stratified in clusters by gender, age, region, and occupation, participated in the CORoNaWork study. We confirmed that very few of the participants belong to the same company. A database of 27,036 individuals was created by excluding 6,051 individuals with invalid responses. As such, we analyzed 13,468 office workers in this database. Manual workers such as assembly-line worker, construction worker, electricians, etc., and hospitality workers such as concierge, waiter/waitress, beautician, etc. were excluded because it was supposed that they might have difficulty in telecommuting and consequently were exempted.

This study was approved by the ethics committee of the University of Occupational and Environmental Health, Japan(reference No. R2-079). Informed consent was obtained in the form of the website.

### Questionnaire

The questionnaire items used in this study were described in detail by Fujino et al (Fujino et al., 2021). The questionnaire collected data on information, including sex, age, educational background, area of participants’ residence, job type, company size of participants’ workplace, working hours per day, family structure, telecommuting frequency, and telecommuting preference. As for telecommuting frequency, we asked participants, “Do you telecommute? Please choose the answer that is closest to your current situation,” and respondents chose one of the following five options: four days a week or more, two to three days a week, one day a week, more than once a month but less than once a week, and never.

Participants were divided into four levels by telecommuting frequency: telecommuting ≥4 days/week (4. high), 2–3 days/week (3. moderate), telecommuting once a week to once a month (2. low), and not-telecommuting (1. none). Regarding telecommuting preference, we asked participants, “How do you feel about telecommuting? (telecommuting preference).” Respondents chose one of five options: “I want to telecommute as much as possible (strongly agree with telecommuting);” “I want to telecommute often (slightly agree with telecommuting);” “either is fine (neither agree nor disagree);” “I want to work in my workplace often (slight disagree with telecommuting);” and “I want to work in my workplace as much as possible (strongly disagree with telecommuting).”

### Cumulative COVID-19 Incidence by Prefecture and Group Classification of Prefectures by Cumulative COVID-19 Incidence

In this study, we used data on the cumulative COVID-19 incidence in each prefecture of Japan. Japan Broadcasting Corporation’s data on cumulative COVID-19 incidence by prefecture (Japan Broadcasting Corporation [NHK], 2020) was downloaded and tabulated from January 15, 2020, when patients with COVID-19 were first identified in Japan, to December 22, 2020, the start date of this study’s questionnaire. Using information from the Statistics Bureau of the Ministry of Internal Affairs and Communications, the cumulative COVID-19 incidence per 100,000 people was calculated using the 2020 population data for each prefecture (Statistics Bureau, 2021); see Supplementary Table 1).

According to the cumulative COVID-19 incidence in each prefecture, 47 prefectures were divided into three levels. High-level regions were composed of 17 prefectures with a cumulative COVID-19 incidence of ≥100 per 100,000 population. Middle-level regions included 19 prefectures with a cumulative COVID-19 incidence of ≥50 per 100,000 population. Finally, low-level regions were composed of 15 prefectures with a cumulative COVID-19 incidence of ≤50 per 100,000 population. As of December 22, 2020, the prefecture with the highest cumulative COVID-19 incidence was Tokyo (371.2/100,000 people), and the lowest was Akita (10.0/100,000 people; Supplementary Table 1). Using data on prefectural residence data, participants living in the high, middle, and low-level regions were assigned to the H, M, and L groups, respectively.

### Variables

We set telecommuting frequency and preference as the outcome variables. The participant groups according to the level of cumulative COVID-19 incidence in each prefecture (H, M, and L group) were set as predictor variables. Sex, age, educational background, job type, company size, working hours per day, marital status, and presence of family living together were adjusted for potential confounders.

### Statistical Analysis

We employed ordinal logistic regression analysis (OLR) to analyze the dependent variables—telecommuting frequency or preference—and the groups (H, M, and L groups). We treated the H, M, and L groups as independent variables and adjusted for sex and age. We also adjusted for multivariate factors, such as personal, work-related, and familial factors.

Cox and Snell R-squared analyses were used to determine the goodness of fit of the statistical model. The p-values of logistic regression analysis were calculated by considering each category scale of the groups as continuous variables (p for trend). In all tests, the threshold for significance was set at p < 0.05. SPSS 25.0 J analytical software (IBM, NY) was used for the statistical analyses.

## Results

### Participants and Descriptive Data

There were 5,641, 4,549, and 3,278 participants, respectively, in the H, M, and L groups (see Figure 1). Participant characteristics classified by telecommuting frequency groups are shown in Table 1. The H group had a higher proportion of participants who were over 50 years old, university graduates or had finished graduate school, managers, and working at companies with more than 10,000 employees. Meanwhile, the L group had a higher proportion of participants who were aged under 40 years, junior or senior high school graduates, and working at companies with 10 to 49 employees. The proportion of participants who telecommuted more than two days per week was 34.7%, 16.3%, and 12.2% in the H, M, and L groups, respectively. The proportion of participants who wanted to telecommute were 48.4%, 36.4%, and 34.3% in the H, M, and L groups, respectively; participants in the H group tending to have higher telecommuting preference. In the L group, more participants wanted to work in their place of employment (36.4%) than telecommute.

**Figure 1.**
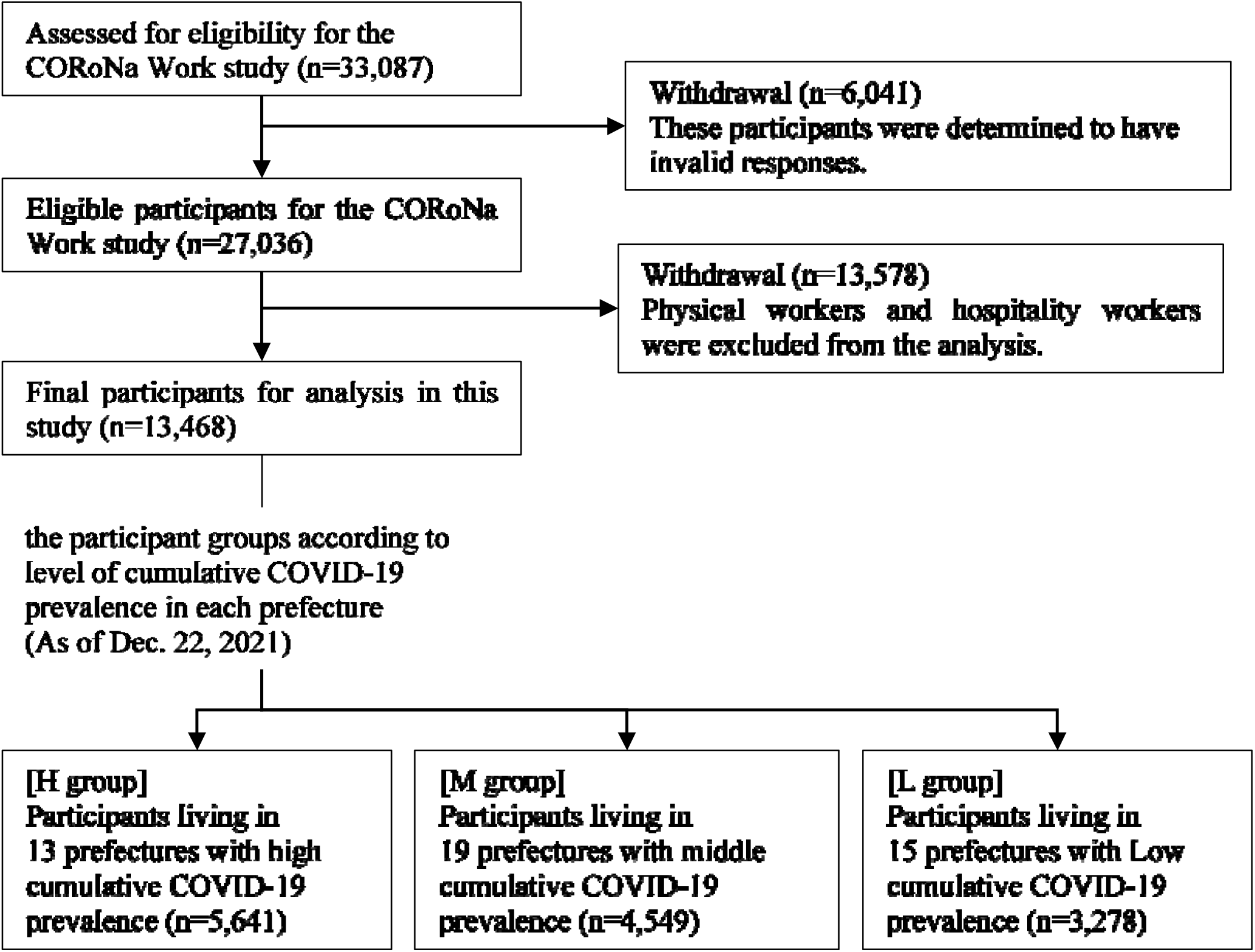
Flow chart of Sample Selection in this Study.

**Table 1.**
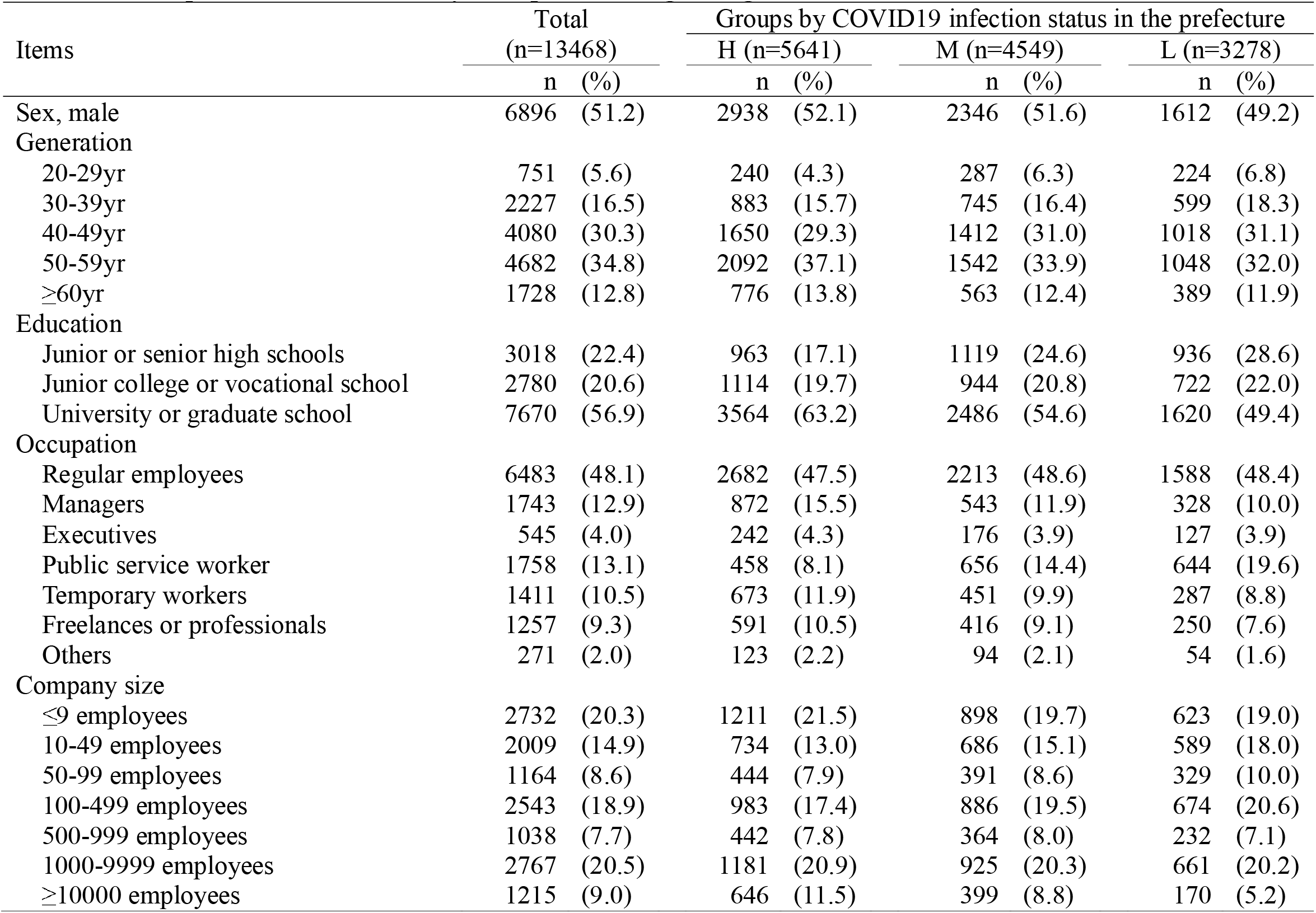
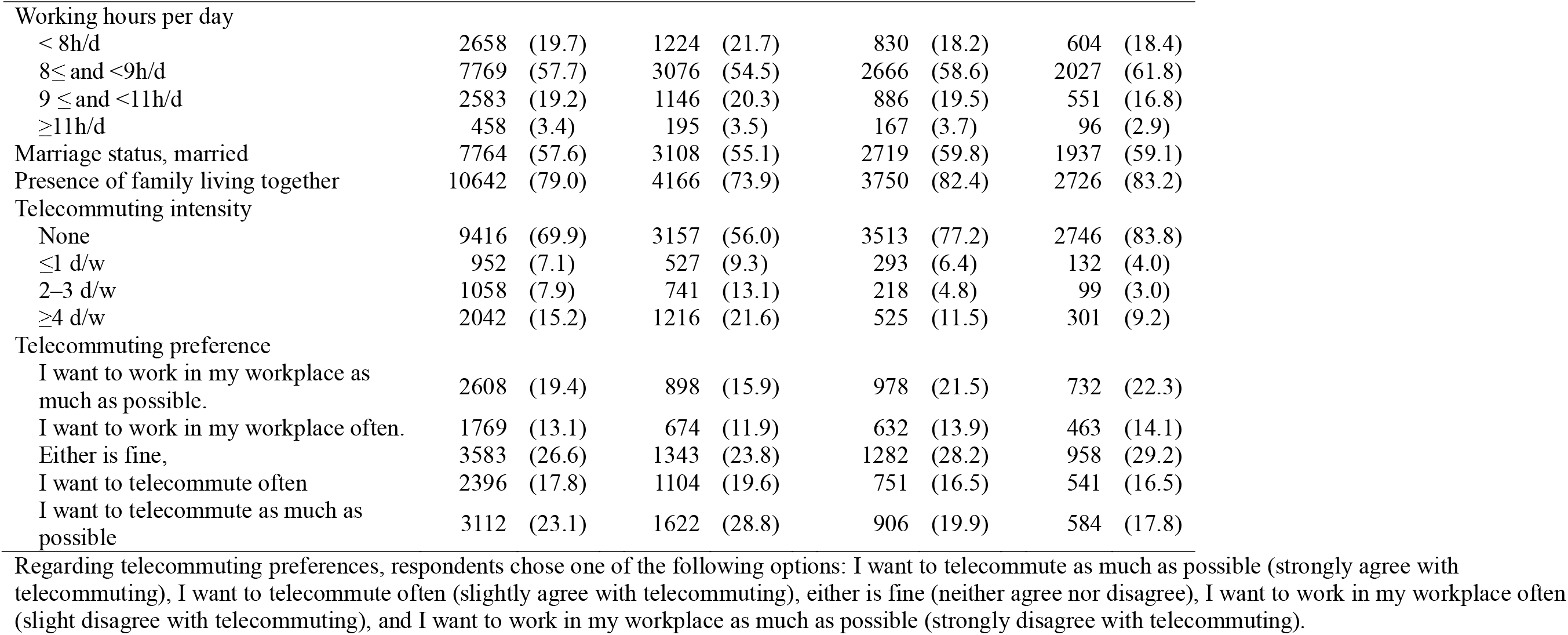
Participants’ Characteristics by Groups According to Regional Cumulative COVID-19 Incidence; n (%)

### Comparison of Telecommuting Frequency and Preference According to the Cumulative COVID-19 Incidence in the Prefectures

Table 2 presents a comparison of telecommuting frequency and preference among groups, according to cumulative COVID-19 incidence in each prefecture. Participants in the H group had higher telecommuting frequency. In sex-age adjusted OLR models, the telecommuting frequency of participants was signficantly higher in the H and M groups compared to the L group (OR=3.83, 95% CI: 3.44–4.25, OR=1.49, 95% CI: 1.33–1.67). In the multivariate adjusted OLR models, the telecommuting frequency of participants was also significantly higher in the H and M groups compared to the L group (OR=3.32, 95% CI: 2.96–3.72, OR=1.29, 95% CI: 1.14–1.47).

**Table 2.**
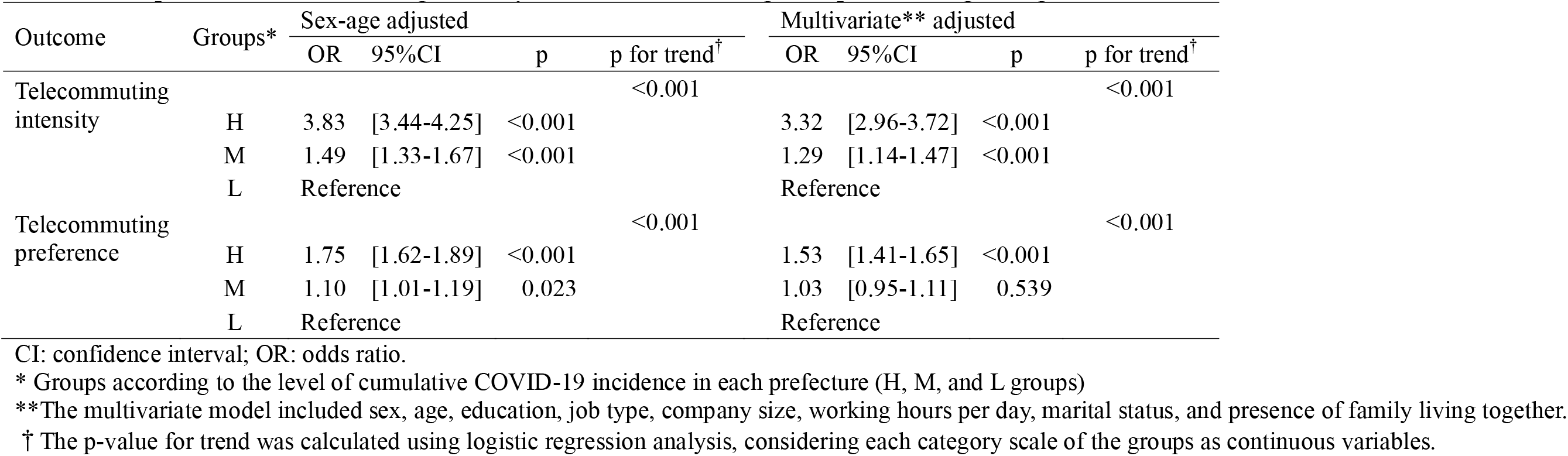
Comparison of Telecommuting Intensity and Preference Among Groups According to Regional Cumulative COVID-19 Incidence.

| Outcome | Groups* | Sex-age adjusted |  |  |  | Multivariate** adjusted |  |  |  |
| --- | --- | --- | --- | --- | --- | --- | --- | --- | --- |
|  |  | OR | 95% CI | p | p for trend<sup>†</sup> | OR | 95% CI | p | p for trend<sup>†</sup> |
| Telecommuting intensity | H | 3.83 | [3.44-4.25] | <0.001 | <0.001 | 3.32 | [2.96-3.72] | <0.001 | <0.001 |
|  | M | 1.49 | [1.33-1.67] | <0.001 |  | 1.29 | [1.14-1.47] | <0.001 |  |
|  | L | Reference |  |  |  | Reference |  |  |  |
| Telecommuting preference | H | 1.75 | [1.62-1.89] | <0.001 | <0.001 | 1.53 | [1.41-1.65] | <0.001 | <0.001 |
|  | M | 1.10 | [1.01-1.19] | 0.023 |  | 1.03 | [0.95-1.11] | 0.539 |  |
|  | L | Reference |  |  |  | Reference |  |  |  |

CI: confidence interval; OR: odds ratio.
\* Groups according to the level of cumulative COVID-19 incidence in each prefecture (H, M, and L groups)
\*\*The multivariate model included sex, age, education, job type, company size, working hours per day, marital status, and presence of family living together.
† The p-value for trend was calculated using logistic regression analysis, considering each category scale of the groups as continuous variables.

In the sex-age adjusted OLR models, telecommuting preference was significantly higher in the H and M groups compared to in the L group (OR=1.75, 95%CI: 1.62–1.89, OR=1.10, 95% CI: 1.01–1.19). However, in the multivariate adjusted ORL model, the telecommuting preference in the M group was not significantly higher compared to L groups (OR=1.03, 95% CI: 0.95–1.11).

## Discussion

This study provides an overview of the telecommuting state based on the cumulative COVID-19 incidence in each prefecture of Japan. We found that as the regional cumulative COVID-19 incidence increased, the telecommuting frequency among office workers also increased. Furthermore, we found that 35% of office workers telecommuted more than two days a week in regions with a high level cumulative COVID-19 incidence, which is approximately 20% higher than in other regions. A survey regarding telecommuting by the Ministry of Land, Infrastructure, Transport, and Tourism (MLIT) reported that the proportion of employees who telecommuted before the COVID-19 pandemic were approximately 9–10% from 2017 to 2019 (Ministry of Land, Infrastructure, Transport and Tourism, 2020). Although the aggregation method in this study differs from that of the survey conducted by the MLIT, it suggests that telecommuting has been widely introduced in the wake of the COVID-19 pandemic.

The current increase in the number of telecommuters could be attributed to the Japanese government’s request to implement methods to control the spread of the COVID-19 pandemic. Therefore, the COVID-19 pandemic may be suppressed depending on telecommuting frequency (Vyas & Butakhieo, 2020; World Health Organization, 2020). However, from January to March 2021, after the survey period in this study, there were many new occurrences of COVID-19 in regions with a high cumulative COVID-19 incidence. A previous study has reported that the average number of contacts decreased by 81% with a regional lockdown in France, and the basic reproductive number (R0), that is the reproduction number when there is no immunity from past exposures or vaccination, nor any deliberate intervention in disease transmission, decreased from 3.18 (95% CI [3.09, 3.24]) before lockdown to 0.68 (95% CI [0.66, 0.69]) during lockdown (Di Domenico, Pullano, Sabbatini, Boëlle, & Colizza, 2020). Given the current state of telecommuting in Japan, it is difficult to drastically reduce the average number of contacts, as has been observed in previous studies. Furthermore, prefectures with a high cumulative COVID-19 incidence have a high population density, and the effect of telecommuting on controlling the COVID-19 epidemic could be limited. When COVID-19 spread nationally, the government was instructed to encourage telecommuting, and the telecommuting frequency increased temporally. However, when instructions were lifted, the telecommuting frequency decreased. Regardless, we speculate that it is difficult to effectively control t telecommuting during the COVID-19 pandemic.

Regarding telecommuting preferences, we observed several trends. The telecommuting preference of participants in regions with a high cumulative COVID-19 incidence tended to be much higher than in other regions, and about half of the participants preferred to telecommute. However, in regions with a low cumulative COVID-19 incidence, the proportion of participants who wanted to telecommute was lower than those who wanted to work from the office. A previous study reported that a high perception of the risk of COVID-19 infection can increase anxiety, and people become more aware of preventive measures for the COVID-19 (Kwok et al., 2020). However, we considered other factors. The great difference between the industrial structures of urban and rural prefectures (Ministry of Health, Labor and Welfare, 2015) may have affected the results of this study. For example, in urban prefectures, the proportion of wholesale/retail and finance/insurance industries is high, while the proportion of factories with production sites is low (Ministry of Health, Labor and Welfare, 2015). In this study, we found that in regions with a high cumulative COVID-19 incidence, there were high proportions of participants who were well-educated, working in large-scale companies, and engaged in managerial positions; this might have positively affected the availability and adaptability of telecommuting. The traffic conditions in each region may also have had an effect. In this study, because regions with a high cumulative COVID-19 incidence were mostly urban prefectures, there might be many opportunities to use public transportation for commuting and travel. It has been reported that riding a crowded train for a long time increases the R0 of influenza (Furuya, 2007). Conversely, in regions with a moderate or low cumulative COVID-19 incidence, it is speculated that many people commute to work by private car. Therefore, if preventive measures for the COVID-19 epidemic are implemented in the workplace, telecommuting preference as a preventive measure may not increase.

This study has certain limitations. Given that the CORoNaWork survey is an internet-based survey, the generalizability of the results is uncertain. To address this issue, this study used cluster sampling—stratified by gender, region, and occupation. On the other hand, since few participants belonged to the same companies, this study can be interpreted as a company-based survey. This study was also a cross-sectional study, and it is unclear how telecommuting or telecommuting preferences varied during the COVID-19 epidemic. We believe that future longitudinal studies may clarify the relationship between the COVID-19 epidemic and telecommuting. Some examples are the sustainability of telecommuting frequency when the COVID-19 epidemic is declining, and the minimum degree of telecommuting frequency required to control the spread of the pandemic.

## Conclusion

In this study, we reviewed the prevalence of telecommuting, based on the cumulative COVID-19 incidence in each prefecture of Japan in the third wave of December 2020. In the region with a high cumulative COVID-19 incidence, 35% of office workers telecommuted more than two days per week. In these regions, both telecommuting frequency and preference were higher than in other areas. Further studies are needed to determine the extent to which telecommuting is effective as a preventive measure for the COVID-19 epidemic and how to promote effective telecommuting.

## Data Availability

Due to the nature of this research, the participants of this study did not consent to their data being publicly shared; hence, supporting data will not be made available.

## Acknowledgements

We appreciate all the participants and members of the CORoNaWork Study Group. The current members of the CORoNaWork Project, in alphabetical order, are as follows: Dr. Yoshihisa Fujino (present chairperson of the study group), Dr. Akira Ogami, Dr. Arisa Harada, Dr. Ayako Hino, Dr. Hajime Ando, Dr. Hisashi Eguchi, Dr. Kazunori Ikegami, Dr. Kei Tokutsu, Dr. Keiji Muramatsu, Dr. Koji Mori, Dr. Kosuke Mafune, Dr. Kyoko Kitagawa, Dr. Masako Nagata, Dr. Mayumi Tsuji, Ms. Ning Liu, Dr. Rie Tanaka, Dr. Ryutaro Matsugaki, Dr. Seiichiro Tateishi, Dr. Shinya Matsuda, Dr. Tomohiro Ishimaru, and Dr. Tomohisa Nagata. All members were affiliated with the University of Occupational and Environmental Health, Japan. We would also like to thank Editage (www.editage.com) for English language editing.

## References

Alon, T. M., Doepke, M., Olmstead-Rumsey, J., & Tertilt, M. (2020). The impact of COVID-19 on gender equality (No. w26947). National Bureau of economic research. Retrieved from https://www.nber.org/system/files/working_papers/w26947/w26947.pdf

Belzunegui-Eraso, A., & Erro-Garcés, A. (2020). Teleworking in the context of the COVID-19 crisis. Sustainability, 12(9), 3662. https://doi.org/10.3390/su12093662

Di Domenico, L., Pullano, G., Sabbatini, C. E., Boëlle, P. Y., & Colizza, V. (2020). Impact of lockdown on COVID-19 epidemic in Île-de-France and possible exit strategies. BMC Med, 18(1), 240. https://doi.org/10.1186/s12916-020-01698-4

Dwivedi, Y. K., Hughes, D. L., Coombs, C., Constantiou, I., Duan, Y., Edwards, J. S., Gupta, B., Lal, B., Misra, S., Prashant, P. (2020). Impact of COVID-19 pandemic on information management research and practice: Transforming education, work and life. International Journal of Information Management, 55, 102211. https://doi.org/10.1016/j.ijinfomgt.2020.102211

Fujino, Y., Ishimaru, T., Eguchi, H., Tsuji, M., Seichiro, T., Ogami, A., Mori K., Matsuda, S. (2021). Protocol for a nationwide Internet-based health survey in workers during the COVID-19 pandemic in 2020. Journal of UOEH. PPR: PPR278797 https://doi.org/10.1101/2021.02.02.21249309

Furuya, H. (2007). Risk of transmission of airborne infection during train commute based on mathematical model. Environmental Health and Preventive Medicine, 12(2), 78–83. https://doi.org/10.1007/BF02898153

Gursoy, D., & Chi, C. G. (2020). Effects of COVID-19 pandemic on hospitality industry: review of the current situations and a research agenda. Journal of Hospitality Marketing and Management, 29, 527–529. https://doi.org/10.1080/19368623.2020.1788231

Japan Broadcasting Corporation (NHK). (2020). Special Site: New Coronavirus. Retrieved from https://www3.nhk.or.jp/news/special/coronavirus/data/

Khan, R., & Javed Hasan, S. (2020). Telecommuting: The problems & challenges during COVID-19 (2020). International Journal of Engineering Research & Technology (IJERT), 9(7), 1027–1033. http://dx.doi.org/10.17577/IJERTV9IS070432

Kwok, K. O., Li, K. K., Chan, H. H. H., Yi, Y. Y., Tang, A., Wei, W. I., & Wong, S. Y. S. (2020). Community responses during early phase of COVID-19 epidemic, Hong Kong. Emerging Infectious Diseases, 26(7), 1575–1579. https://doi.org/10.3201/eid2607.200500

Lai, C.C., Shih, T.P., Ko, W.C., Tang, H.J., & Hsueh, P.R. (2020). Severe acute respiratory syndrome coronavirus 2 (SARS-CoV-2) and corona virus disease-2019 (COVID-19): The epidemic and the challenges. International Journal of Antimicrobial Agents, 55(3), 105924. https://doi.org/10.1016/j.ijantimicag.2020.105924

Ministry of Health, Labour and Welfare, Japan. (2015). Analysis of the labor economy, 2015 edition. Retrieved from https://www.mhlw.go.jp/wp/hakusyo/roudou/15/15-1.html

Ministry of Health, Labour and Welfare, Japan. (2020). Decisions by the headquarters for novel coronavirus disease control. Basic policies for novel coronavirus disease control. Retrieved from https://www.mhlw.go.jp/stf/seisakunitsuite/bunya/newpage_00032.html

Ministry of Land, Infrastructure, Transport and Tourism. (2020). FY 2020 Telework Population Survey. Retrieved from https://www.mlit.go.jp/toshi/daisei/content/001392107.pdf

Persol Research and Consulting Co., Ltd. (2020a). The first and second urgent survey on the impact of countermeasures against a new coronavirus on Telework. 2020. Retrieved from https://rc.persol-group.co.jp/thinktank/research/activity/data/telework.html

Persol Research and Consulting Co., Ltd. (2020b). The fourth urgent survey on the impact of countermeasures against a new coronavirus on Telework. Retrieved from https://rc.persol-group.co.jp/thinktank/research/activity/data/telework-survey4.html

Persol Research and Consulting Co., Ltd. (2020c). The third urgent survey on the impact of countermeasures against a new coronavirus on Telework. 2020. Retrieved from https://rc.persol-group.co.jp/thinktank/research/activity/data/telework-survey3.html

Statistics Bureau, Ministry of Internal Affairs and Communications, Japan. (2021). Statistics. Retrieved from http://www.stat.go.jp/english/index.html

Tokyo Metropolitan Government. (2021). Results of a survey on telework adoption rates. Retrieved from https://www.metro.tokyo.lg.jp/tosei/hodohappyo/press/2021/03/06/14.html

Vyas, L., & Butakhieo, N. (2020). The impact of working from home during COVID-19 on work and life domains: an exploratory study on Hong Kong. Policy Design and Practice, 1–18. https://doi.org/10.1080/25741292.2020.1863560

World Health Organization. (2020). Getting your workplace ready for COVID-19. Retrieved from www.google.com/url?sa=t&rct=j&q=&esrc=s&source=web&cd=&cad=rja&uact=8&ved=2ahUKEwiVo5L41L7wAhXW7GEKHdDvCnIQFjAAegQIBBAD&url= www.who.int%2Fdocs%2Fdefault-source%2Fcoronaviruse%2Fgetting-workplace-ready-for-covid-19.pdf&usg=AOvVaw1lyz1pQ5J7LqKSxuwW4bsp

