## Supplementary Table 1. for "Telecommuting frequency and preference among Japanese workers according to regional cumulative COVID-19 incidence: a cross-sectional study"

Supplementary Table 1. Classification by Level of Cumulative COVID-19 Incidence in Each Prefecture and Cumulative COVID-19 Incidence in Each Prefecture

| Level of cumulative incidence of COVID-19 in each prefecture | Prefectures | Population (×1,000 people) | Cumulative incidence of COVID-19 per 100,000 population  as of Dec. 22, 2021 |
| --- | --- | --- | --- |
| High level regions | Tokyo | 13921 | 371.2 |
|  | Okinawa | 1453 | 341.9 |
|  | Osaka | 8809 | 309.1 |
|  | Hokkaido | 5250 | 236.0 |
|  | Aichi | 7552 | 188.8 |
|  | Kanagawa | 9198 | 184.9 |
|  | Saitama | 7350 | 160.5 |
|  | Hyogo | 5466 | 151.6 |
|  | Kyoto | 2583 | 149.1 |
|  | Fukuoka | 5104 | 146.1 |
|  | Chiba | 6259 | 145.8 |
|  | Nara | 1330 | 124.5 |
|  | Gunma | 1942 | 101.2 |
| Middle level regions | Gifu | 1987 | 88.5 |
|  | Gunma | 1138 | 85.6 |
|  | Hiroshima | 2804 | 85.1 |
|  | Kumamoto | 1748 | 84.7 |
|  | Hiroshima | 2306 | 77.7 |
|  | Ibaraki | 2860 | 75.3 |
|  | Kochi | 698 | 75.1 |
|  | Shiga | 1414 | 66.8 |
|  | Shizuoka | 3644 | 65.8 |
|  | Mie | 1781 | 65.1 |
|  | Wakayama | 925 | 63.7 |
|  | Miyazaki | 1073 | 62.8 |
|  | Yamanashi | 811 | 60.5 |
|  | Okayama | 1890 | 59.0 |
|  | Kagoshima | 1602 | 55.5 |
|  | Tochigi | 1934 | 53.8 |
|  | Nagano | 2049 | 51.6 |
|  | Oita | 1135 | 51.2 |
|  | Saga | 815 | 50.8 |
| Low level regions | Toyama | 1044 | 48.6 |
|  | Fukui | 768 | 44.2 |
|  | Fukushima | 1846 | 41.3 |
|  | Yamaguchi | 1358 | 34.9 |
|  | Nagasaki | 1327 | 32.0 |
|  | Aomori | 1246 | 31.8 |
|  | Yamagata | 1078 | 30.4 |
|  | Ehime | 1339 | 28.3 |
|  | Iwate | 1227 | 27.7 |
|  | Shimane | 674 | 27.1 |
|  | Tokushima | 728 | 26.6 |
|  | Kagawa | 956 | 22.0 |
|  | Niigata | 2223 | 20.1 |
|  | Tottori | 556 | 12.7 |
|  | Akita | 13921 | 10.0 |
